# The Effect of Frames on COVID-19 Vaccine Hesitancy

**DOI:** 10.1101/2021.01.04.21249241

**Authors:** Risa Palm, Toby Bolsen, Justin T. Kingsland

## Abstract

In order to control the spread of infectious diseases such as COVID-19, it will be important to develop a communication strategy to counteract “vaccine hesitancy”. This paper reports the results of a survey experiment testing the impacts of several types of message content: the safety and efficacy of the vaccine itself, the likelihood that others will take the vaccine, and the possible role of politics in promoting the vaccine. In an original survey of 1123 American M-Turk respondents, we provided six different information conditions suggesting the safety and efficacy of the vaccine, the lack of safety/efficacy of the vaccine, the suggestion that most others would take the vaccine, the suggestion that most others would not take the vaccine, the suggestion that the vaccine is being promoted to gain greater control over individual freedom, and the suggestion that it is being rushed for political motivations. We compared the responses for those in the treatment groups with a control group who received no additional information. In comparison to the control group, those who received information about the safety/efficacy of the vaccine were more likely to report that they would take the vaccine, those who received information that others were reluctant to take the vaccine were more likely to report that they themselves would not take it, that other Americans would not take it, and that it was not important to get the vaccine, and those who received information about political influences on vaccine development expressed hesitancy to take it. Communication of effective messages about the vaccine will be essential for public health agencies that seek to promote vaccine take-up.

## Introduction

Vaccination programs have reduced the toll of infectious diseases by preventing infection or reducing the severity of symptoms, contributing to higher standards of pubic health by reducing morbitiy and mortality rates (Andre et al., 2008). But vaccination programs can only be effective when they are accepted by large segments of the population. Vaccine hesitancy, the reluctanct of patients to receive vaccines, has been identified by the World Health Organization as one of the top ten threats to global health (Puri et al., 2020; World Health Organization, 2019). In the COVID-19 pandemic of 2020-2, as in future outbreaks of vaccine-preventable illness, it will be important to combat this hesitancy and to promote the uptake of the vaccine through effective communication strategies (Nyhan et al., 2014; French et al., 2020). In order to develop a communication strategy that is effective, an understanding of how different types of information may influence the public’s beliefs and vaccination intentions will be required. The contribution of this paper is the examination of the casual effect of exposure to distinct pro- and anti-vaccination message frames on individuals’ intentions to get vaccinated. The case study we examine here is the 2020 situation surrounding beliefs about a vaccine for COVID-19, but the implications are transferrable to the acceptance of vaccines developed in the future as well.

## Framing effects and vaccination beliefs

Communication about the development and testing of any vaccine is transmitted through “frames” used in any message. For example, a *media frame, or frame in communication*, refers to “words, images, phrases, and presentation styles that a speaker (e.g., a politician, a media outlet) uses when relaying information about an issue or event to an audience” (Chong & Druckman, 2007, p. 100). An *emphasis framing effect* occurs when exposure to a media frame causes an audience to privilege the specific consideration(s) made salient when forming an overall opinion on any issue due to increases in the accessibility and/or perceived strength of the frame(s) (Druckman, 2001). We focus exclusively on *emphasis framing effects* and not *equivalency framing effects* that occur when positive or negative information unconsciously influences preferences (Tversky & Kahneman, 1981; Druckman, 2004). For example, a communicator might emphasize the personal or the public health benefits of practicing physical distancing to combat COVID-19 thereby increasing an audience’s willingness to follow the recommended behavior (Goldberg et al., 2020; Jordan et al., 2020).

In a review of 316 articles on framing in health communication, Guenther et al. (2020) noted that most experimental studies to date have evaluated the relative persuasiveness of “gain” as opposed to “loss” frames, creating a gap in the research literature in more fully understanding the effects of thematic frames (see also Penta, & Baban, 2018). The empirical study described here contributes to the understanding of the impact of message framing on vaccine uptake. We tested the impacts of several types of emphasis frames: two emphasizing the safety and efficacy (or their absence) of the vaccine, two emphasizing the likelihood that taking the vaccine would be in accord (or not) with general social norms, one suggesting that the entire discussion of vaccines is being shaped by those who want to exert more government control over individual behavior, and one suggesting that any rush to develop a vaccine in order to meet political deadlines could result in a less than optimal vaccine. Each of these frames was expected to result in specific changes in attitude about the vaccine.

### Perceptions about the safety and efficacy of a vaccine

Because of its significance to public health, there have been numerous studies of the factors that cause “vaccine hesitancy” (Hornsey et al., 2018; Puri et al., 2020; Thunström et al., 2020). Most of the studies have focused on decision-making in the context of parents vaccinating their children, the acceptance of the HPV vaccine, or decision-making with respect to the flu vaccine (Brewer et al., 2007; Callaghan et al., 2020; Dubé et al., 2013; Kim et al., 2019; Nan et al., 2015; Smith et al., 2017).

The influence of perceived safety on vaccine hesitancy has been a finding of several meta-analyses of the scientific literature. In a review of 2791 studies published between 1990-2019, Sweileh (2020) found that although the reasons for vaccine hesitancy varied depending on the disease and on the cultural and national context, the overwhelming reason for hesitancy was fears about the safety of the vaccines. Yaqub et al. (2014) reviewed 1187 articles published between 2009 and 2012, primarily about HPV and flu vaccines, and found that “fear of adverse side effects and vaccine safety” were the leading reasons for hesitancy, both in the general population and among healthcare professionals. Similarly, a review of 2895 articles in English, French and Spanish from 2004-2014 (Karafillakis & Larson, 2017, p. 4846) found that although different concerns were expressed about vaccine safety for different types of vaccines, the “largest area of concern was vaccine safety.” In a study of childhood vaccine safety, van der Linden et al. (2015) found that agreement with a statement that “90% of medical scientists agree that vaccines are safe” was the most important predictor of public support for vaccines. Similarly, an analysis of 25 national samples from 12 different countries showed that “trust in experts” was the most consistent predictor of vaccine acceptance (Kerr et al., 2020). Finally, a more recent study found that presenting individuals with information specifically about a COVID-19 vaccine’s safety increased Americans’ plans to get vaccinated (Motta, 2020).

Vaccine efficacy has also been identified as a separate and important dimension of the decision-process (Motta, 2020). In a study manipulating an H1N1 vaccination message along with perceived safety, efficacy, susceptibility to the disease and severity, Nan et al (2012) found that the most important factor in the acceptance of the vaccine among older adults was perceived efficacy. Similarly, Chapman and Coups (1999) found that perceived efficacy of the flu vaccine was the most important factor in its acceptance by healthy adults, followed closely by the likelihood that it would not have side effects.

In the case of the COVID-19 vaccine, based on the large body of empirical literature emphasizing the importance of both safety and efficacy in the decision to accept a vaccine, we propose the following: *Individuals presented with a message that emphasizes the safety and effectiveness (or lack of safety and ineffectiveness) of a vaccination for COVID-19 will shift their beliefs and behavioral intentions in the direction of the message* (**Hypothesis 1**).

### Perceptions about the intentions of others

A long scholarly tradition has demonstrated the impact of “social norms” on behavior changes. Social norms are the “tacit rules that members of a group implicitly recognize and that affect their decisions and behavior” (Brewer et al., 2020, p.170). Cialdini et al (1991) distinguished two distinct types of social norms: those that are “injunctive”, informing people about what is approved or disapproved and those that are “descriptive” of typical or common behavior. Examples of experimental manipulations of social norms to change behavior include studies of college binge drinking (Perkins & Craig, 2002), smoking (Linkenbach & Perkins, 2003), hotel towel reuse (Goldstein et al., 2008), and energy conservation (Schultz et al., 2007).

The underlying principle for the impact of descriptive social norms is that most people want to bring their behavior in line with what they perceive to be the behavior of others. Brewer et al (2017, p. 171) suggest that “because individuals learn about descriptive social norms, in part, by observing the behavior others, it should be possible to change social norms by sufficiently changing the behavior of the group”. A descriptive social norms marketing campaign will therefore outline what a majority of people are doing in order to get the target audience to conform.

Research has shown that “strategic messaging” of the social norms can have an influence on behavior decisions ranging from voting (Gerber et al., 2008) to using weight-loss products (Lim et al., 2020) or charitable giving (Croson, et al., 2009). Indeed, the application of the scholarly findings about social norms into popular marketing has been familiar in the multitude of advertisements that suggest that “everyone else” is buying or participating in what is being sold (Melnyk et al., 2019).

Social norms may also have a negative effect on behavior because the perception that “everyone is *not* doing it” will decrease the intention to act (Kahan, 2014, p. 4). In such instances, communicating a descriptive social norm may result in unintended effects: providing information about what people are neglecting to do may result in fewer people engaging in the pro-social behavior that is being promoted (Murray & Matland, 2014, Rimal & Real, 2005; but see Hassell & Wyler, 2019).

Several studies have examined the impact of social norms on the adoption of various vaccines (Xiao & Borah, 2020). Allen et al (2009) found that social norms, that is, the perceived behavior of friends who either had already been vaccinated or were considering the vaccine, were the strongest predictors of the intent to be vaccinated against human papillomavirus (HPV). Brunson (2013) identified the role of descriptive social norms in parental decisions about their children’s vaccinations. De Bruin et al. (2019) documented the impact of perceived vaccine coverage in the social circle (defined as people with whom the respondent had regular contact) on vaccination behavior for influenza. Similarly, Parker et al (2013) found that social influence, that is, the likelihood that people around the respondent were being vaccinated, was the most common reason for choosing to get a flu vaccine.

Because of the consensus in the literature concerning the likelihood that people will try to make their behavior conform with their perceptions of the behavior of others, we hypothesize that: *Exposure to a message that emphasizes other Americans’ willingness (or unwillingness) to get vaccinated for COVID-19 will shift their beliefs and behavioral intentions in the direction of the message* (**Hypothesis 2**).

### Perceptions about the role of politics

Vaccine hesitancy not only has a long history, but it also reflects “historical events and individual belief systems reflective of different societal periods “(McAteer et al. 2020, p. 703). Not surprisingly, then, the issues around the development of a COVID-19 vaccine too have become an issue intertwined with politics in U.S. A content analysis of newspaper and television coverage surrounding the issue from March to May 2020 showed that politicians are featured as often or more often than scientists (Hart, Chinn, & Soroka, 2020). A recent survey in the US found that an increase in conservatism also increased the odds of vaccine hesitancy; moreover, those who intended to vote for President Trump in 2020 were 35% more likely to report that they would refuse a COVID-19 vaccination (Callaghan et al., 2020). Another study reported that when Republicans were exposed to an anti-vaccination argument posted on Twitter by President Trump, they became more concerned about getting vaccinated (Hornsey et al., 2020).

While conservatives have expressed higher levels of hesitancy toward a COVID-19 vaccine, President Trump stated publicly that approval of a vaccine before November would help his chances for re-election (Irfan, 2020). This frequently stated goal may have contributed further to hesitancy, this time on the part of Democrats, by promoting the belief that scientists were being pressured to approve a vaccine before it had been thoroughly tested for political goals (Cohen, 2020).

A distinct political argument that surfaced in U.S. media was that the government should require “compulsory vaccinations” for COVID-19 “to win the war against the novel coronavirus” (Lederman et al., 2020). This rhetoric sometimes appeared alongside claims that “Operation Warp Speed” was a way to further regulate the lives of Americans and enrich drug companies. We anticipate that exposure to such information reduces individuals’ willingness to get vaccinated We thus predicted that: *Exposure to a message that emphasizes the role that politics is playing in driving the approval of a COVID-19 vaccination will increase vaccine hesitancy*. (**Hypothesis 3**)

## Survey Experiment

We implemented a survey-experiment in August 2020 in which we randomly assigned 1,123 respondents, recruited from Amazon’s Mechanical Turk (MTurk), to one of seven experimental conditions. MTurk is an online crowdsourcing platform commonly used in the social sciences to estimate causal relationships; the results are comparable to identical studies fielded on general population samples (Levay et al., 2016; Mullinix et al., 2015).

Respondents randomly assigned to the *control* condition (*N*=157) were not exposed to any information prior to answering our key outcome measures (described below). Respondents randomly assigned to all other conditions were exposed to a message that varied the headline and content of a short “article” formatted to mimic a news story about a COVID-19 vaccine. We used information from published news articles as the basis for our treatments, although we edited them for length and reading level. We restricted the sample to U.S. respondents who had successfully completed at least 100 tasks and had at least a 95% approval rating on MTurk. The median completion time for the survey was 6.3 minutes and participants received $0.15 in remuneration upon completion. The sample was large and diverse with respect to demographic and political characteristics: for instance, 41% of respondents identified as Republicans, 22% identified as Independents, and 37% identified as Democrats. Further, our sample is 55% female and 45% male. Other descriptive statistics for the sample are available in the appendix table A3.

### Experimental Treatments and Conditions

Participants in all conditions completed an IRB-approved consent form and were informed that they would be asked some questions about their opinions related to a COVID-19 vaccine. To complete the survey, respondents had to check a box to indicate they had read the following debriefing statement: “At present there is no FDA-licensed vaccine to prevent COVID-19. Vaccines have been highly effective in preventing a range of serious infectious diseases. The FDA has the scientific expertise to evaluate any potential COVID-19 vaccine candidate regardless of the technology used to produce or to administer the vaccine. This includes the different technologies such as DNA, RNA, protein and viral vectored vaccines being developed by commercial vaccine manufacturers and other entities. For factual information about the regulation of COVID-19 vaccine development, please consult this website from the Food and Drug Administration.” We also provided a link to the FDA website from which this language was drawn.

Respondents randomly assigned to the ***safe and effective*** condition (*N*=172) were presented with the headline, “Scientists Are Working on a Safe and Effective COVID-19 Vaccine”, followed by information that a vaccine would be “safe, have few side effects, and most of all, will be effective in preventing the illness” and that it will have been “carefully tested and evaluated by scientists and medical professionals”. Respondents assigned to the ***unsafe and ineffective*** condition (*N*=159) were presented with the headline, “A COVID-19 Vaccine is Neither Effective nor Safe”, followed by an information calling into question the efficacy of any FDA-approved vaccine by noting that it will be approved by the FDA if it shows “only 50% efficacy”, suggesting that it could have serious side-effects, and that immunity could last only for a few months. Respondents in the ***willing*** condition (*N*=171) were presented with the headline, “Most American Say They Will Get Vaccinated against COVID-19”, followed by information that included the results from “a recent tracking survey” that “indicated widespread willingness in the U.S.” to take the vaccine. The treatment included additional details explaining why most Americans are willing to get vaccinated. Conversely, respondents in the ***unwilling*** condition (*N*=157) saw the headline, “Many Americans Say They Will Not Get Vaccinated against COVID-19”, followed by information from a “recent tracking survey” that “indicated widespread reluctance in the U.S. to take any COVID-19 vaccination”. It included additional details explaining why many Americans may be hesitant to take the vaccine. Two additional conditions invoked “politics” in an anti-vaccination message. In the ***agenda*** condition (*N*=149), respondents were presented with the headline, “Liberal Media Pushing Agenda for ‘Mandatory Vaccinations’ and ‘Immunization Cards’’, followed by information suggesting that the rush to develop a COVID-19 vaccine is a way to enrich pharmaceutical companies and for the government to assert greater control over the lives of individuals. In the ***Trump*** condition (*N*=158), respondents read the headline, “President Trump Pushing for Rapid Approval of a Covid-19 Vaccine”, followed by information raising concern that the FDA might approve a vaccine due to political pressure prior to Election Day, as an “October surprise”. Full wording for these treatments is presented in Appendix Table A1.

### Dependent Measures

Participants in all conditions responded to three key outcome measures post-treatment. First, we asked respondents, “If an FDA-approved vaccine against the coronavirus becomes widely available, how likely is it that **you** will get vaccinated?” (1=extremely unlikely; 7=extremely likely). Second, we asked respondents, “How likely is it that ***other Americans*** will get vaccinated if an FDA-approved vaccine against the coronavirus becomes widely available?” (1=extremely unlikely; 7=extremely likely). Third, we asked respondents, “In general, how ***important*** do you believe it is that all Americans get vaccinated for the coronavirus once an FDA-approved vaccine is widely available?” (1=not at all important; 7=extremely important).

## Results

To test our hypotheses, we estimate OLS regression models with robust standard errors. We regress each dependent variable on our condition indicators, omitting the *Control* condition as the reference group. In all models, cell entries contain OLS coefficients representing the difference in means between the treatment condition and the *control* condition. We also included a manipulation check at the end of the survey where respondents in the treatment conditions were asked if the “article” they read earlier was opposed to or supportive of getting vaccinated for COVID-19. The treatments were accurately perceived in the directions we intended across all conditions (Appendix Table A2).

### Intentions to Take the Vaccine

Our first hypothesis was that one-sided frames highlighting considerations related to the safety and effectiveness of a COVID-19 vaccine would shift intentions to take the vaccine in the direction of the message. As we predicted (H1), respondents who read the *safe and effective* treatment were more likely to express an intention to get vaccinated (*b = 0*.*36, p=0*.*05*, column 1, Table 1). However, this effect is driven exclusively by the response among Independents (*b = 0*.*84, p=0*.*04*, column 3) and Democrats (*b = 0*.*78, p=0*.*01*, column 4). Counter to our prediction (H1), reading the *not safe* treatment had no effect on respondents’ willingness to get vaccinated. Our second hypothesis was that one-sided frames highlighting a descriptive social norm would influence individuals’ beliefs and behavioral intentions. As we predicted (H2), the *unwilling* treatment decreased reported intentions to take a COVID-19 vaccine (*b = −0*.*40, p=0*.*04*, column 1, Table 1), whereas the *willing* treatment increased reported intentions to get vaccinated (*b = 0*.*43, p=0*.*03*). The effect of the norm-based treatments appear to have exerted similar effects across partisan in terms of the “direction” of the treatment’s impact, although the frames again exerted the strongest effect on Independents and Democrats. Our third hypothesis was that exposure to a message emphasizing the role of politics would decrease intentions to get vaccinated. We found no main effect of either *agenda* or *Trump* on personal behavioral intentions.

**Table 1.**
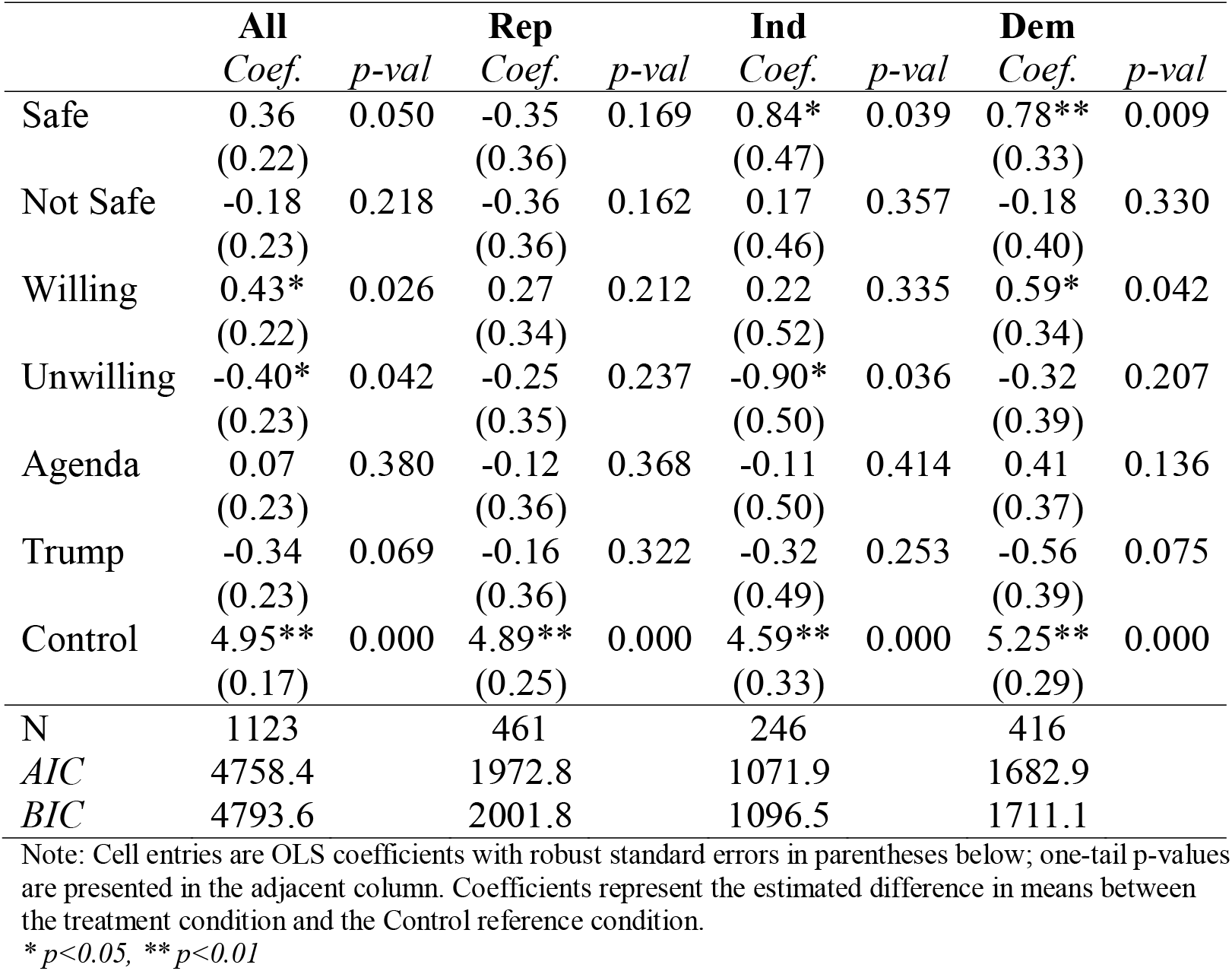
Will Take Vaccine - Treatment Effects.

### Belief that other Americans will take COVID-19 vaccine

Our second dependent variable was the belief that other Americans will take a COVID-19 vaccine. First, we find mixed support for Hypothesis 1. While there was no main effect of the *safe and effective* message, the *not safe* treatment reduced perceptions that other Americans will take the vaccine (*b = −0*.*38, p=0*.*01*, column 1, Table 2). The direction of the *not safe* message’s effect was consistent across partisan subgroups, but had the strongest impact on Independents (*b = −0*.*61, p=0*.*04*). We also find mixed support for Hypothesis 2. Although the *willing* message had no main effect on perceptions about other Americans’ willingness to take the vaccine, exposure to the *unwilling* treatment reduced the belief that other Americans will get vaccinated (*b = −1*.*06, p=0*.*00*). This effect was significant across all partisan subgroups in the sample relative to their counterparts in the control condition. As we predicted (H3), exposure to the political messages had a negative effect on respondents’ beliefs about whether other Americans will take the vaccine, with the *agenda* and *Trump* treatments moving respondents in those conditions approximately one-quarter point and one-half point on the response scale, respectively ((*b = −0*.*27, p=0*.*04; b = −0*.*45, p=0*.*00*).

**Table 2.** Believe Other Americans Take COVID-19 Vaccine.

|  | **All** |  | **Rep** |  | **Ind** |  | **Dem** |  |
| --- | --- | --- | --- | --- | --- | --- | --- | --- |
|  | *Coef.* | *p-val* | *Coef.* | *p-val* | *Coef.* | *p-val* | *Coef.* | *p-val* |
| Safe | -0.00 | 0.499 | -0.39 | 0.051 | 0.30 | 0.181 | 0.23 | 0.148 |
|  | (0.14) |  | (0.23) |  | (0.33) |  | (0.22) |  |
| Not Safe | -0.38** | 0.009 | -0.28 | 0.113 | -0.61* | 0.036 | -0.40 | 0.072 |
|  | (0.16) |  | (0.23) |  | (0.34) |  | (0.27) |  |
| Willing | 0.21 | 0.067 | -0.03 | 0.445 | 0.41 | 0.081 | 0.35 | 0.060 |
|  | (0.14) |  | (0.23) |  | (0.29) |  | (0.23) |  |
| Unwilling | -1.06** | 0.000 | -0.88** | 0.000 | -1.29** | 0.000 | -1.16** | 0.000 |
|  | (0.16) |  | (0.24) |  | (0.37) |  | (0.26) |  |
| Agenda | -0.27* | 0.036 | -0.20 | 0.185 | -0.28 | 0.182 | -0.35 | 0.091 |
|  | (0.15) |  | (0.23) |  | (0.31) |  | (0.26) |  |
| Trump | -0.45** | 0.002 | -0.35 | 0.063 | -0.65* | 0.037 | -0.40 | 0.055 |
|  | (0.16) |  | (0.23) |  | (0.36) |  | (0.25) |  |
| Control | 5.34** | 0.000 | 5.55** | 0.000 | 5.14** | 0.000 | 5.23** | 0.000 |
|  | (0.10) |  | (0.15) |  | (0.23) |  | (0.17) |  |
| N | 1122 |  | 461 |  | 245 |  | 416 |  |
| AIC | 3973.7 |  | 1645.9 |  | 882.0 |  | 1441.1 |  |
| BIC | 4008.9 |  | 1674.8 |  | 906.5 |  | 1469.3 |  |

Note: Cell entries are OLS coefficients with robust standard errors in parentheses below; one-tail p-values are presented in the adjacent column. Coefficients represent the estimated difference in means between the treatment condition and the Control reference condition.
\* $p < 0.05$ , \*\* $p < 0.01$

### Belief it is Important for All to Get Vaccinated

The experimental treatments also had an effect on respondents’ belief that getting vaccinated for COVID-19 is important for all Americans. Counter to our expectation (H1), there was no effect of exposure to the *safe and effective* or the *not safe* treatments on this outcome measure. In fact, we observe an unexpected *negative* impact of the *safe and effective* message on Republicans’ beliefs about the importance of getting the vaccine (*b = −0*.*49, p=0*.*05*, column 2, Table 3). As we predicted (H2), the *unwilling* treatment decreased perceptions of the importance of taking a COVID-19 vaccine (*b = −0*.*57, p=0*.*00*), whereas the *willing* treatment increased perceptions about its importance (*b = 0*.*32, p=0*.*03*). Also as we predicted (H3), the *agenda* message decreased perceptions about the importance of taking a COVID-19 vaccine (*b = −0*.*37, p=0*.*03*); however, the effect of the *Trump* condition fell below conventional levels of statistical significance to reject the null hypothesis. We again observe the largest negative impact of the political messages on Independents in the sample.

**Table 3.** Important All Get COVID-19 Vaccine.

|  | **All** |  | **Rep** |  | **Ind** |  | **Dem** |  |
| --- | --- | --- | --- | --- | --- | --- | --- | --- |
|  | *Coef.* | *p-val* | *Coef.* | *p-val* | *Coef.* | *p-val* | *Coef.* | *p-val* |
| Safe | 0.03<br>(0.18) | 0.425 | -0.49*<br>(0.30) | 0.049 | 0.41<br>(0.38) | 0.147 | 0.32<br>(0.25) | 0.099 |
| Not Safe | -0.21<br>(0.19) | 0.126 | -0.06<br>(0.29) | 0.422 | -0.33<br>(0.40) | 0.204 | -0.31<br>(0.30) | 0.153 |
| Willing | 0.32*<br>(0.17) | 0.028 | 0.46*<br>(0.25) | 0.035 | -0.00<br>(0.41) | 0.496 | 0.28<br>(0.27) | 0.144 |
| Unwilling | -0.57**<br>(0.19) | 0.002 | -0.42<br>(0.31) | 0.084 | -1.43**<br>(0.42) | 0.000 | -0.24<br>(0.28) | 0.194 |
| Agenda | -0.37*<br>(0.19) | 0.027 | -0.42<br>(0.28) | 0.069 | -0.84*<br>(0.43) | 0.024 | -0.00<br>(0.30) | 0.500 |
| Trump | -0.28<br>(0.18) | 0.060 | -0.12<br>(0.29) | 0.334 | -0.59<br>(0.41) | 0.072 | -0.27<br>(0.27) | 0.159 |
| Control | 5.34**<br>(0.13) | 0.000 | 5.14**<br>(0.19) | 0.000 | 5.22**<br>(0.27) | 0.000 | 5.66**<br>(0.22) | 0.000 |
| N | 1123 |  | 461 |  | 246 |  | 416 |  |
| AIC | 4317.3 |  | 1807.2 |  | 985.8 |  | 1448.2 |  |
| BIC | 4352.4 |  | 1836.1 |  | 1010.4 |  | 1476.4 |  |

Note: Cell entries are OLS coefficients with robust standard errors in parentheses below; one-tail p-values are presented in the adjacent column. Coefficients represent the estimated difference in means between the treatment condition and the Control reference condition.
\* $p < 0.05$ , \*\* $p < 0.01$

## Conclusion / Discussion

It is crucial to know how messages the public receives about a novel vaccine ultimately shape decisions regarding whether or not to get vaccinated (Mheidly & Fares, 2020). The results we report demonstrate that exposure to messages about a vaccine can have a powerful influence on intentions to be immunized, beliefs about whether other Americans will get vaccinated, and assessments of the importance of this collective action to combat the spread of a virus. In this study, information that highlighted the safety and efficacy of an approved vaccine against COVID-19, or that called into question its safety and effectiveness, influenced related perceptions and beliefs. The results also demonstrate the powerful impact that communicating descriptive social norms can exert on beliefs and behavioral intentions. Messages emphasing descriptive social norms, especially those which highlighted the hesistancy of other Americans to take a COVID-19 vaccine, had a strong effect on vaccine perceptions and intentions, and this effect was observed across partisan groups in our sample. The experimental treatments that accentuated the role of politics in driving decisions about a COVID-19 vaccine also shaped beliefs and intentions.

In order to combat vaccine hesitancy, it is urgent that messaging be carefully and thoughtfully crafted, taking into account what social scientists have learned about the factors that influence message acceptance. This study suggests that, given the specific historical setting in which the Covid-19 virus spread in 2020-21, messages will be mostly likely to succeed if they emphasize safety and efficacy, use positive social norms, and stress the independence of the development of the vaccine from political influence. These are likely to be essential elements of any successful messaging campaign to increase a vaccine’s uptake.

## Data Availability

Data are available upon request.

## Notes

### Competing Interest Statement

The authors have declared no competing interest.

### Clinical Trial

No clinical trials were performed

### Funding Statement

No external funding

### Author Declarations

Georgia State University Institutional Review Board (IRB) Exempt Protocol Category 2 Reference Number 361974 Approved August 27, 2020

